# Clinical and laboratory features of COVID-19 illness and outcomes in immunocompromised individuals during the first pandemic wave in Sydney, Australia

**DOI:** 10.1101/2023.08.17.23293358

**Authors:** Nila J. Dharan, Sarah C. Sasson, Golo Ahlenstiel, Christopher R. Andersen, Mark Bloch, Griselda Buckland, Nada Hamad, Win Min Han, Anthony D. Kelleher, Georgina V. Long, Gail V. Matthews, Michael M. Mina, Emmanuelle Papot, Kathy Petoumenos, Sanjay Swaminathan, Barbara Withers, James Yun, Mark N. Polizzotto, the CORIA Study Group

## Abstract

People with immunocompromising conditions are at increased risk of SARS-CoV-2 infection and mortality, however early in the pandemic it was challenging to collate data on this heterogenous population. We conducted a registry study of immunocompromised individuals with polymerase chain reaction (PCR)-confirmed SARS-CoV-2 infection from March – October 2020 in Sydney, Australia to understand clinical and laboratory outcomes in this population prior to the emergence of the Delta variant. 27 participants were enrolled into the study including people with a haematologic oncologic conditions (n=12), secondary immunosuppression (N=8) and those with primary or acquired immunodeficiency (i.e. HIV; N=7). All participants had symptomatic COVID-19 with the most common features being cough (64%), fever (52%) and headache (40%). Five patients demonstrated delayed SARS-CoV-2 clearance lasting three weeks to three months. The mortality rate in this study was 7% compared to 1.3% in the state of New South Wales Australia during the same period. This study provides data from the first eight months of the pandemic on COVID-19 outcomes in at-risk patient groups.

## Introduction

Early in the COVID-19 pandemic, male sex, advanced age and certain comorbidities were identified as risk factors for death following SARS-CoV-2 infection [1-4]. Large international studies with data on immunocompromising conditions identified that patients with haematological malignancies, connective tissue disease, solid organ transplantation and those on immunosuppressive therapies for other indications were also at increased risk of death [4]. A study of 310 immunocompromised individuals with SARS-CoV-2 infection in the United Kingdom reported that, compared with the general population, mortality among hospitalised immunocompromised individuals was greater (38% vs. 26%) and immunocompromised individuals were younger at time of death [5]. In Australia, chronic immunosuppression was also identified to be associated with increased risk of death in patients in intensive care units early in the pandemic [6].

The Coronavirus Outcomes Registries in Immunocompromised Individuals Australia (CORIA) was a clinical registry study of adults with immunocompromising conditions and SARS-CoV-2 infection acquired between March – October 2020, prior to COVID-19 vaccines, specific therapies and the emergence of variants of concern (VOCs). The aim of this study was to determine the clinical and laboratory COVID-19 outcomes of adult patients with immunocompromising conditions.

## Methods

The study included individuals with specified immunocompromising conditions aged 18 years or older with polymerase chain reaction (PCR)-confirmed SARS-CoV-2 infection between March 1, 2020 – October 31, 2020. Participants had been tested after developing symptoms consistent with COVID-19 and/or being identified as a close contact of a confirmed case. Participants were enrolled from nine clinical sites in Sydney, Australia: seven major teaching hospitals with high caseloads of immunocompromised individuals; one primary care centre with a high caseload of people with HIV; and one specialist melanoma centre. Eligible immunocompromising conditions included: primary immunodeficiency, defined as a predisposition to infection associated with a genetic or inherited deficit of immune function; current receipt of immunosuppressive therapy (excluding inhaled corticosteroids); HIV infection; undergoing active management for cancer diagnosed within 36 months of enrolment (excluding superficial basal cell and squamous cell carcinomas; including treatment with immune checkpoint inhibitors); and solid organ transplantation. All eligible cases from the nine enrolling sites were included.

All data were sourced from the participants’ medical record. At enrolment, the following data were collected: demographic and clinical characteristics; epidemiologic risk factors for infection; medical history; presenting clinical symptoms; clinical status; COVID-19 treatments; and clinical test results including SARS-CoV-2 and respiratory pathogen PCR testing, haematology, hepatic and renal chemistry, and inflammatory biomarkers. Follow-up data on clinical symptoms, clinical status, laboratory results and treatment history were collected on Days 3, 7, 14, 21, 28 and 3 months.

Three main groups of immunocompromising conditions were categorised as: 1) haematologic and oncologic conditions; 2) primary or acquired immunodeficiency (i.e. HIV); and 3) secondary immunosuppression (including immunosuppressive therapy and solid organ transplantation). The time to end of clinical isolation was defined as the time from initial SARS-CoV-2 positive PCR to either a negative SARS-CoV-2 PCR or resolution of all symptoms after a certain timepoint according to local guidance at the time [7]. Descriptive statistics were used to calculate the proportions of participants with various clinical and laboratory characteristics at presentation and follow up, overall and across immunocompromising conditions group.

The study was reviewed by the St Vincent’s Hospital Human Research Ethics Committee and granted a waiver of the need for individual consent for data collection as per the National Statement on Ethical Conduct in Human Research. The study was registered at Clinicaltrials.gov (NCT04354818).

## Results

Of the 27 participants enrolled, 58% were male and the median age was 66 years. Twelve (44%) of the participants had haematologic (n=7) or oncologic conditions (n=5), 8 (30%) had secondary immunosuppression and 7 (26%) had primary or acquired immunodeficiency (i.e. HIV), as shown in Table 1 (further detail in Supplemental Table 1). Of the total cohort with available data, 41% had a history of hypertension, 26% had diabetes and 19% had cardiovascular disease. Among the 25 participants with clinical symptom data, all had symptomatic disease with the most common presenting symptoms being cough (64%), fever (52%) and headache (40%) (Table 1). Two patients, both in the haematology/oncology group, received treatments for COVID-19 as part of clinical trials (remdesivir n=1; hydroxychloroquine n=1).

**Table 1.**
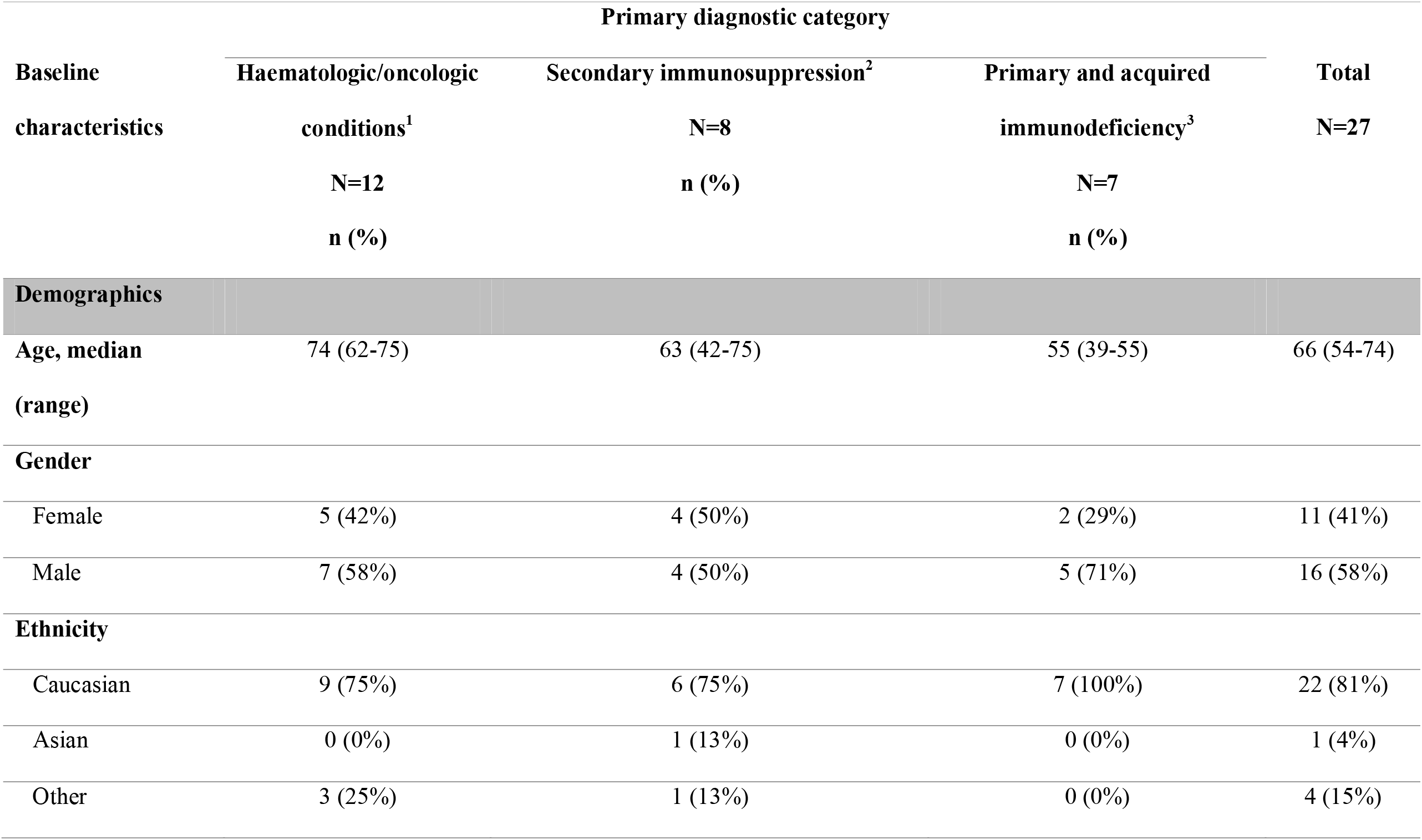
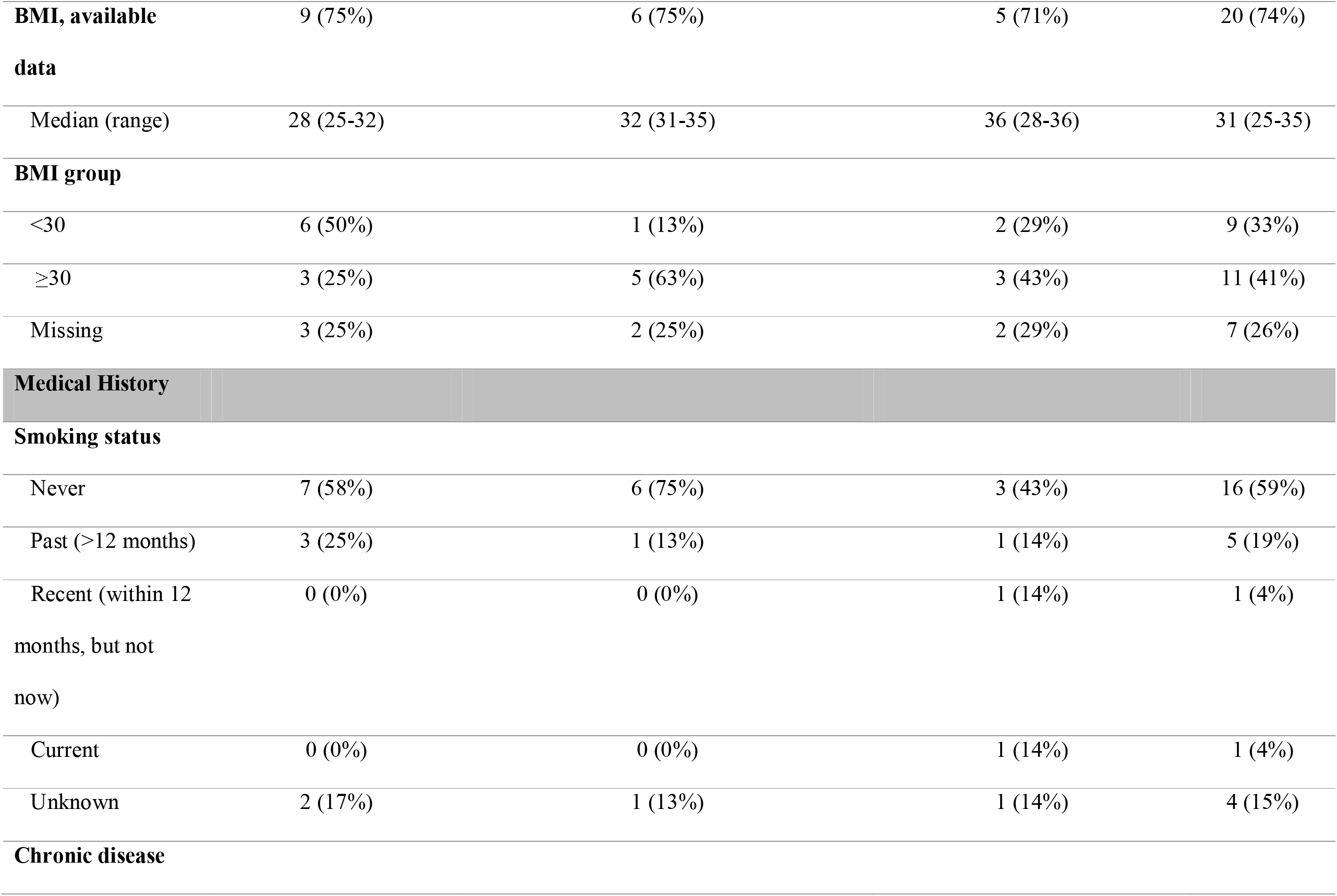
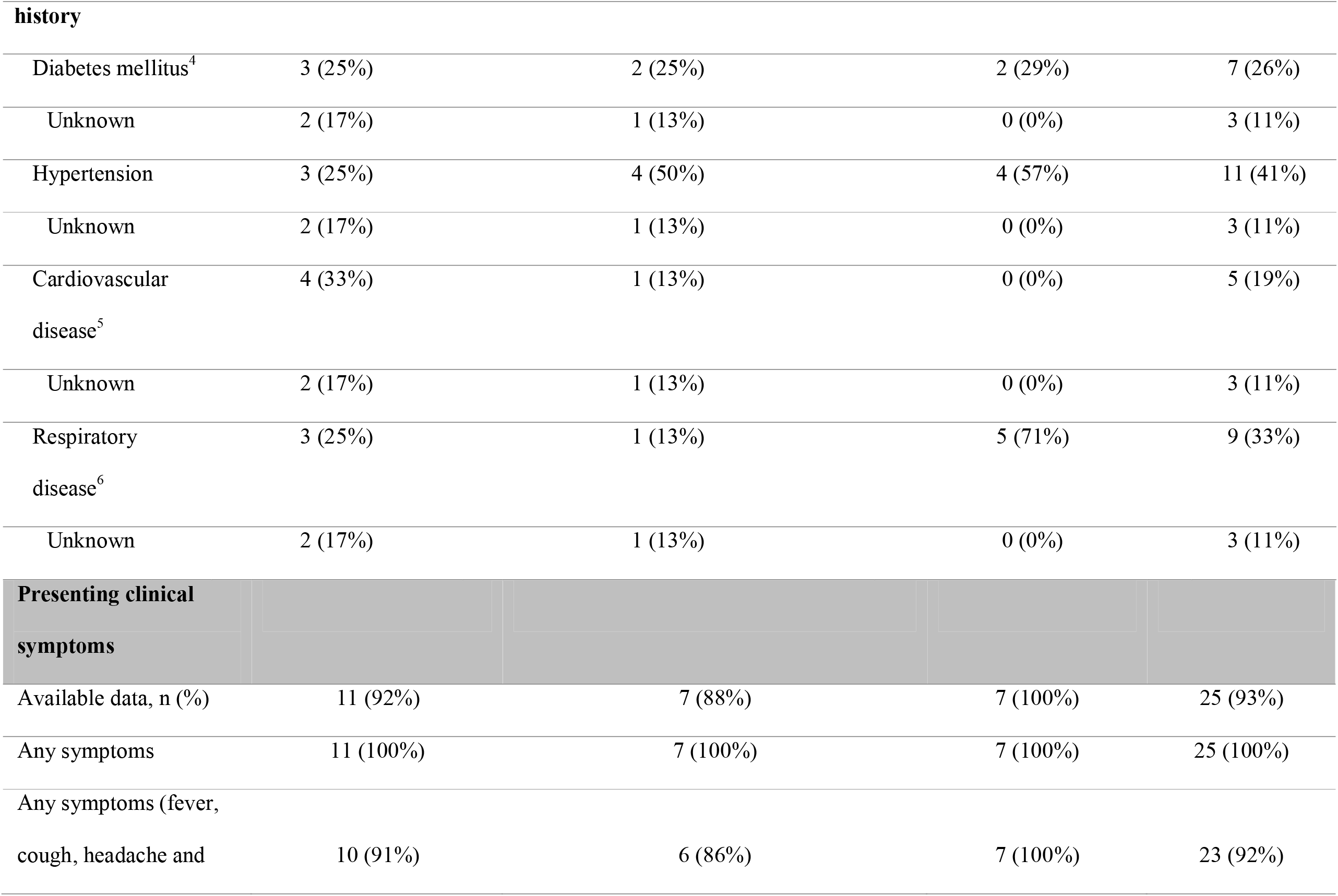
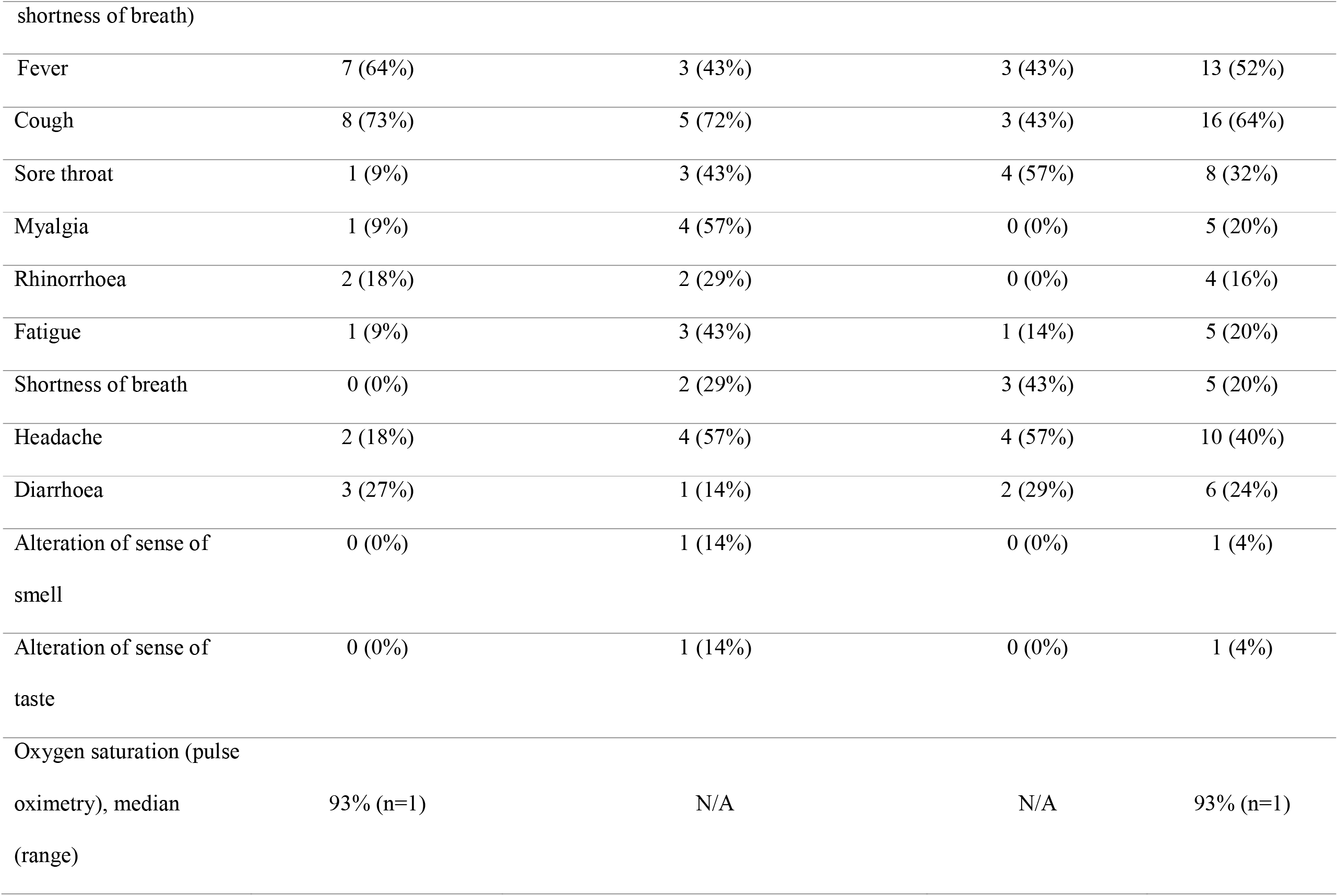
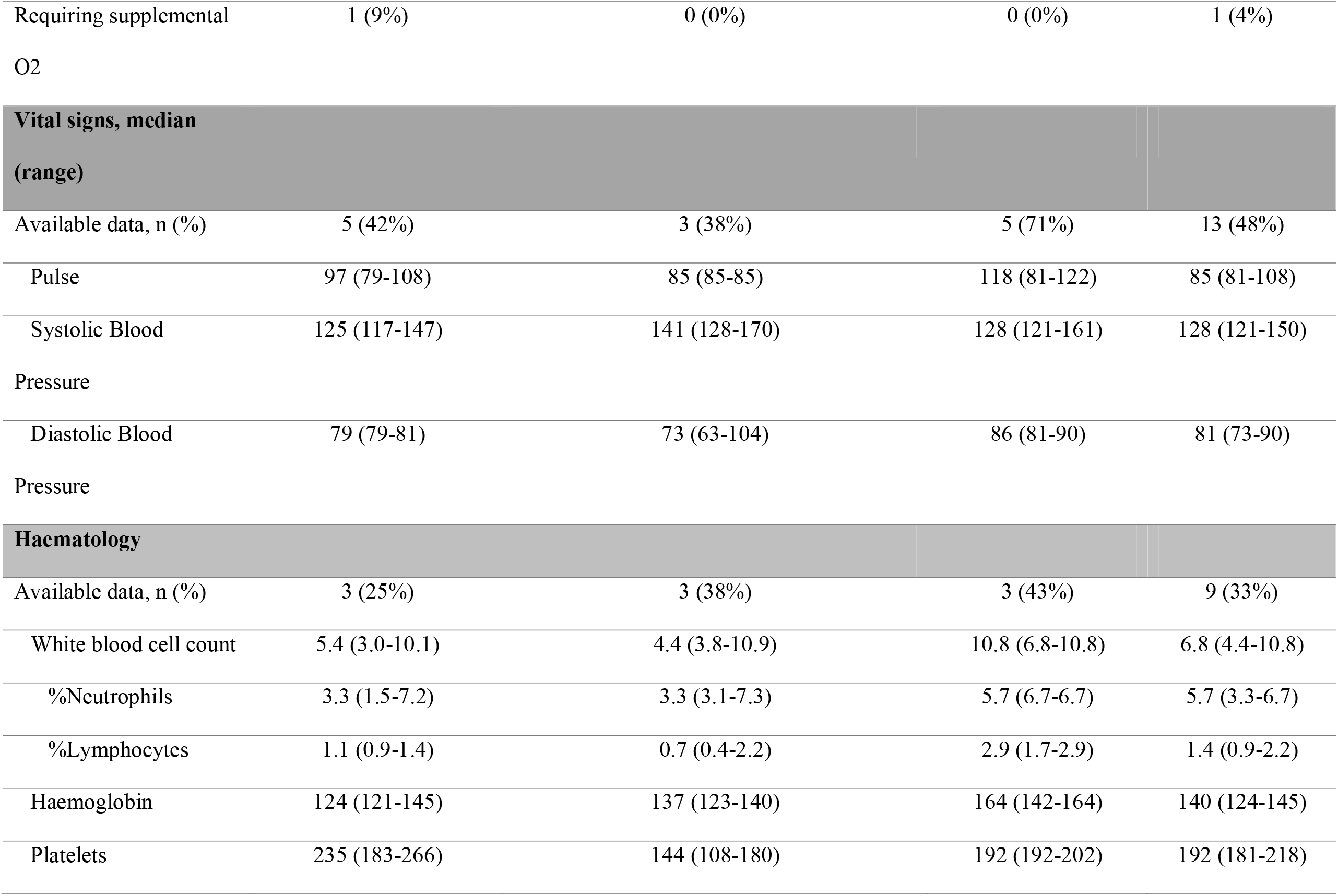
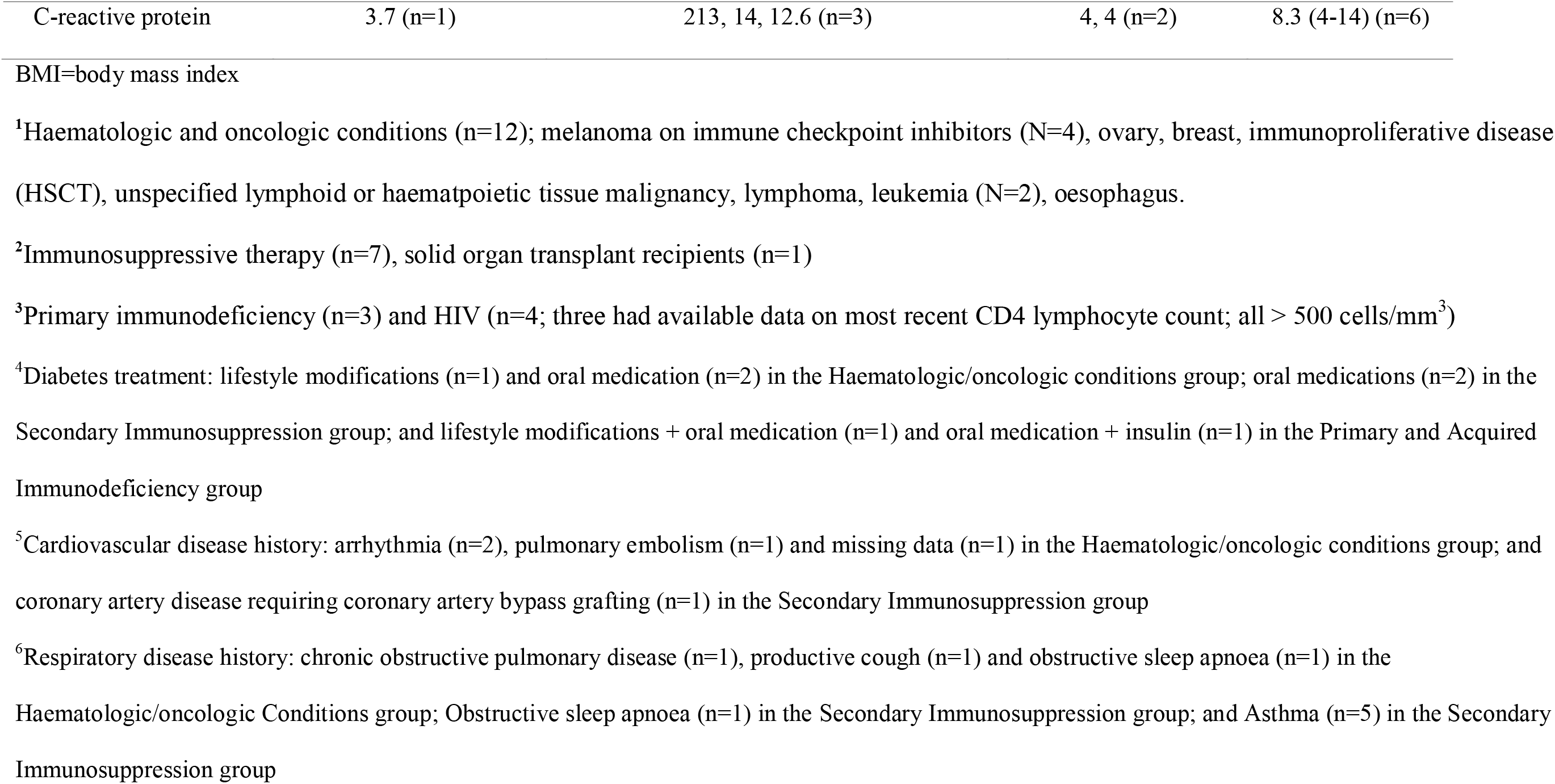
Demographic characteristics, medical history, clinical symptoms and laboratory results at time of presentation, by primary immunocompromising diagnostic category.

At Days 7 and 28, 47% and 22% of participants, respectively, were still reporting symptoms (Figure 1). The median (interquartile range [IQR]) time to release from isolation for the total cohort was 16 (15-27) days: 15 (11-29) days for the haematologic/oncologic group, 26 (17-27) days for the secondary immunosuppression group, and 14 (14-14) days for the primary/acquired immunodeficiency group. Nine (33%) of participants were tested for SARS-CoV-2 by PCR after their entry date test (“Day 0”; see Supplemental Table 2). Of these nine participants, three (33%) tested positive at Day three, one (11%) at Day seven, two (22%) at Day 21, one (11%) at Day 28 and two (22%) at three months. Of the two participants that tested positive at three months, one had melanoma and one had an autoinflammatory condition.

**Figure legend.** Proportion of participants reporting symptoms of SARS-CoV-2 infection over time, by primary immunocompromising diagnostic category. Panel A. Fever; Panel B. Cough; Panel C. Headache; Panel D. Shortness of breath. E. Any symptom

Where available, laboratory results were recorded over three months (Supplemental Figure 1). The platelet count peaked at Day 14, while the neutrophil and lymphocyte counts were highest at baseline and the C-reactive protein was highest at Day three.

Clinical illness outcomes are shown in Supplemental Table 3. Among the total cohort, nine (33%; n=3 from each group) required hospitalisation with the median time from enrolment to hospital admission of three days. Three (11%) participants (two in the secondary immunosuppression group and one in the hematological/oncological group) required mechanical ventilation with the median time from enrolment to initiation of mechanical ventilation of three days. Overall, two (7%) participants died; one was a female in her 60s in the hematological/oncological group and the other was a female in her 80s in the secondary immunosuppression group. The time from SARS-CoV-2 diagnosis to death for both participants was 90 days; both died primarily from their underlying disease, rather than severe COVID-19.

## Discussion

We report demographic and clinical characteristics of individuals with conditions associated with immune compromise who were diagnosed with COVID-19 from March – October 2020 prior to the introduction of SARS-CoV-2 vaccination, COVID-19 treatments, and the emergence of VOCs. During this time, there were few data available on the clinical outcomes of patients with COVID-19, particularly in rare and orphan disease groups, and there was considerable concern that, similar to other respiratory viruses [8-13], immunosuppressed individuals may be at risk for increased risk of mortality associated with SARS-CoV-2 infection and slower viral clearance. All patients in our study had symptomatic illness as well as evidence of prolonged durations of viral shedding.

The most common clinical presenting symptoms among all participants were cough, and fever, which were of similar prevalence to that reported in another cohort of 55 patients (9% were immunocompromised) studied during a similar period in Melbourne, Australia [14] (cough 64% in our cohort vs. 66%; fever 52% in our cohort versus 47%). Other symptoms were also similar including sore throat (32% in our cohort vs. 29%) and diarrhoea (24% in our cohort vs. 29%), although headache was more common among our cohort (40% in our cohort vs. 0.05%).

There is documented evidence of prolonged SARS-CoV-2 shedding in immunocompromised people, which can increase the risk of outgrowth of novel viral variants. The median time to de-isolation was 16 days in our cohort which is comparable to data published in the general population during the pre-Delta phase of the pandemic (median 12-16 days [15, 16]. In our study, the secondary immunosuppression group numerically had the longest median time to viral clearance (26 days) and there was delayed viral clearance in five patients ranging from three weeks to three months, in line with similar previous reports in immunocompromised individuals [17, 18]. A recent case series of SARS-CoV-2 Delta strain infection in immunocompromised individuals has demonstrated that immunocompromised patients can shed infectious (culture-positive) virus for up to 33 days accompanied by low levels of neutralising antibody [17]. These findings have implications for the risk of emergence of new variants with resistance to SARS-CoV-2 therapeutics, as well as for infection control. Additionally, for patients requiring allogeneic hematopoietic stem cell transplantation, chimeric antigen T cell therapies or even chemotherapy, this delayed clearance can delay therapy and potentially negatively a patient’s cancer survival outcomes.

Our findings suggest that patients in the secondary immunosuppression group may be functionally more immunocompromised than the haematological/oncology and primary/acquired immunodeficency groups. This may reflect the fact that four of the 12 patients in the heamatological/oncology group were on immunostimulatory checkpoint inhibitor therapies, which have been associated with similar mortality rates to patients with cancer [19]. In addition, three of the seven participants in the primary/acquired immunodeficiency group were people with HIV on antiretroviral therapy with CD4 lymphocyte counts >500 cells/mm^3^.

In this cohort of immunocompromised individuals, 33% required hospitalisation, 11% required mechanical ventilation, and 7% died. At the end of October 2020 in New South Wales (the end of our study period), among the total 4,237 reported COVID-19 cases, 55 (1.3%) had died [20].

## Conclusions

Our data provide insights into COVID-19 illness and outcomes in immunocompromised individuals in a resource-rich setting prior to COVID-19 vaccines and specific therapies, and at a time when community transmission and healthcare strain was relatively low. These data provide a benchmark for future work evaluating vulnerable populations including those with heterogenous conditions affecting immune system function. We acknowledge several limitations including the small sample size, which reflects the small and short peak of COVID-19 in New South Wales in 2020, and limited data collection such as serial SARS CoV-2 PCR results. Further understanding around which specific subgroups of immunocompromised individuals are at greatest risk of SARS-CoV-2 infection, severe COVID-19 illness outcomes and prolonged SARS-CoV-2 shedding, particularly in the post-vaccination era and with the emergence of new variants, remains an important area of research.

## Data Availability

The datasets from this study are available from the corresponding author upon reasonable request.

## Acknowledgements

#CORIA Study Group: Nila J. Dharan, Sarah C. Sasson, Golo Ahlenstiel, Christopher R. Andersen, Mark Bloch, Griselda Buckland, Nada Hamad, Win Min Han, Anthony D. Kelleher, Georgina V. Long, Gail Matthews, Michael M. Mina, Emmanuella Papot, Kathy Petoumenos, Tri Phan, Sanjay Swaminathan, Barbara Withers, James Yun, Mark N. Polizzotto, David A. Brown, Rowena A. Bull, Matthew S. Carlino, Jennifer Curnow, Sarah Davidson, Dominic E. Dwyer, Prudence N. Gatt, Yuvaraj Ghodke, Sally Hough, Peter MacDonald, Susan Maddocks, Marianne Martinello, Deborah Marriott, Alexander M. Menzies, Tania Sorrell.

**Supplemental Figure Legend**. Markers of infection and inflammation over time among the total cohort (N=27)

## Disclaimer

This publication was funded by the Australian Government Department of Health. The views expressed in this publication do not necessarily represent the position of the Australian Government.

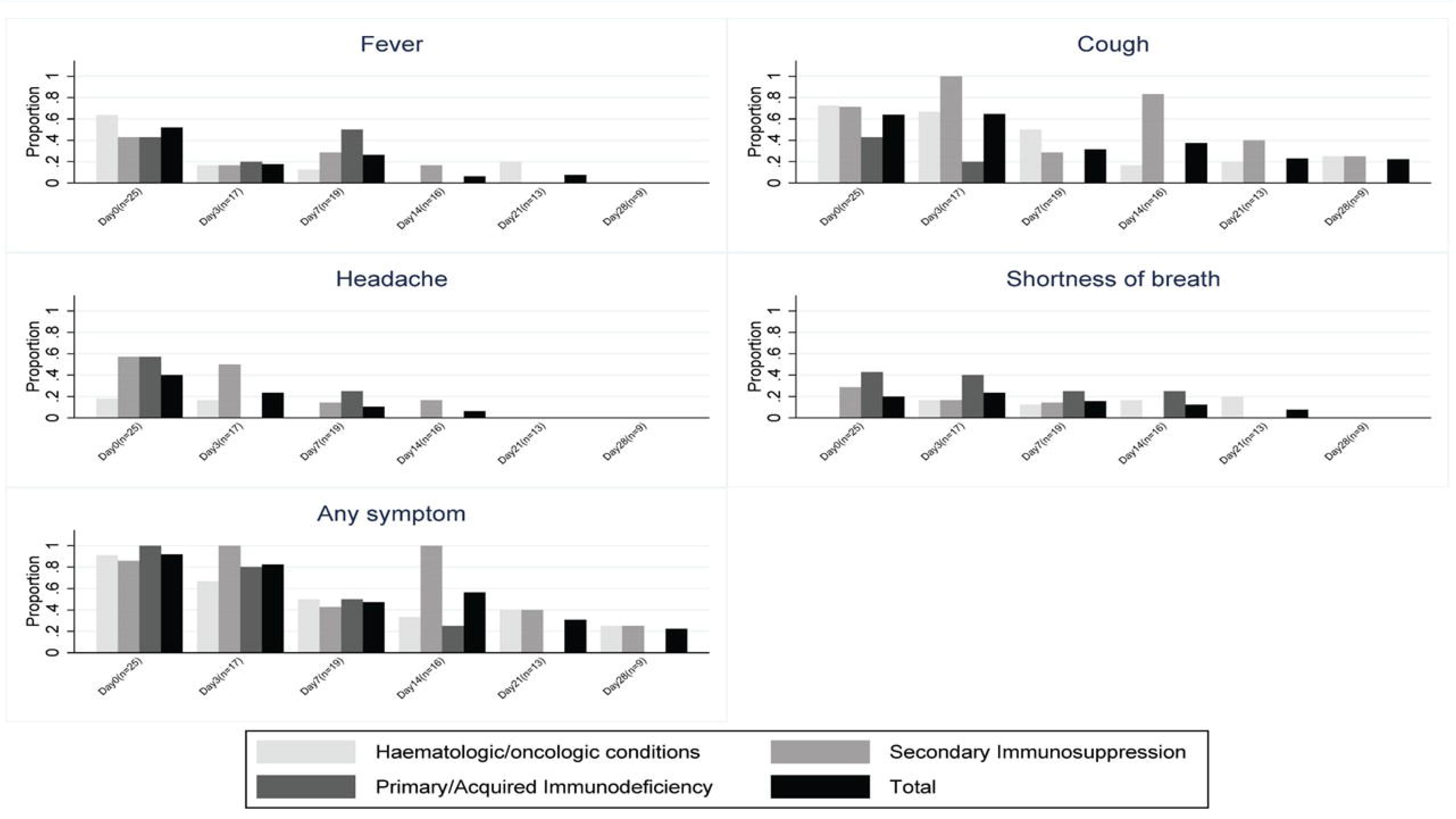

**Supplemental Table 1.** **Medication list** (data available for 19 participants)

| **Participant** | **Primary diagnostic criteria** | **Underlying immunosuppressive condition** | **Medications** |
| --- | --- | --- | --- |
| 1 | Haematologic/oncologic conditions | Cutaneous melanoma | Anti-PD-1, Anti-PDL1 |
| 2 | Haematologic/oncologic conditions | Cutaneous melanoma | Anti-PD-1, Anti-CTLA-4 |
| 3 | Haematologic/oncologic conditions | Cutaneous melanoma | Anti-PD-1, Anti-CTLA-4 |
| 4 | Haematologic/oncologic conditions | Follicular lymphoma | Rituximab, Intravenous immunoglobulin (IvIg) |
| 5 | Haematologic/oncologic conditions | Refractory myeloma | Methylprednisolone |
| 6 | Haematologic/oncologic conditions | Malignant neoplasm of other connective and soft tissue | Anti-PD-1 |
| 7 | Haematologic/oncologic conditions | Malignant immunoproliferative diseases | Ruxolitinib |
| 8 | Haematologic/oncologic conditions | Chronic transfusion dependent myelofibrosis | Ruxolitinib |
| 9 | Haematologic/oncologic conditions | Malignant neoplasm of oesophagus | Trastuzumab (Herceptin), Capecitabine |
| 10 | Primary/acquired immunodeficiency | HIV infection | Elvitegravir/cobicistat/emtricitabine/tenofovir alafenamide |
| 11 | Primary/acquired immunodeficiency | HIV infection | Dolutegravir+darunivir/cobicistat |
| 12 | Primary/acquired immunodeficiency | HIV infection | Dolutegravir+darunivir/cobicistat |
| 13 | Primary/acquired immunodeficiency | HIV infection | Not on HIV treatment |
| 14 | Primary/acquired immunodeficiency | Unknown | Mycophenolate Mofetil, Prednisone |
| 15 | Primary/acquired immunodeficiency | Autoinflammatory Disorders | Methotrexate |
| 16 | Primary/acquired immunodeficiency | Autoinflammatory Disorders | Etanercept, Prednisone |
| 17 | Secondary immunosuppression | Psoriasis | Secukinumab |
| 18 | Secondary immunosuppression | Multiple sclerosis | Fingolimod |
| 19 | Secondary immunosuppression | Rheumatoid arthritis | Methotrexate, Sulfasalazine |
| 20 | Secondary immunosuppression | MOG Antibody Demyelinating Syndrome | Tecifidera, Ocrelizumab |

**Supplemental Table 2.**
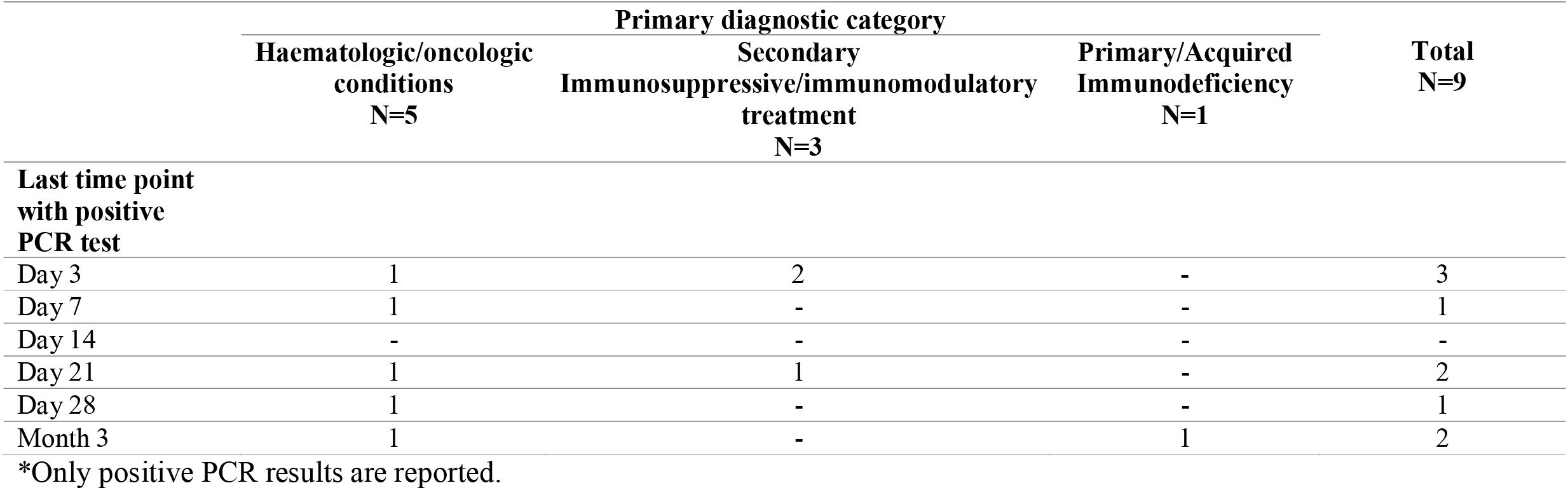
Number of participants with positive COVID-19 PCR positive results over time, by primary immunocompromising diagnostic category*

|  | Primary diagnostic category |  |  | Total<br>N=9 |
| --- | --- | --- | --- | --- |
|  | Haematologic/oncologic<br>conditions<br>N=5 | Secondary<br>Immunosuppressive/immunomodulatory<br>treatment<br>N=3 | Primary/Acquired<br>Immunodeficiency<br>N=1 |  |
| **Last time point**<br>**with positive**<br>**PCR test** |  |  |  |  |
| Day 3 | 1 | 2 | - | 3 |
| Day 7 | 1 | - | - | 1 |
| Day 14 | - | - | - | - |
| Day 21 | 1 | 1 | - | 2 |
| Day 28 | 1 | - | - | 1 |
| Month 3 | 1 | - | 1 | 2 |

\*Only positive PCR results are reported.

**Supplemental Table 3.**
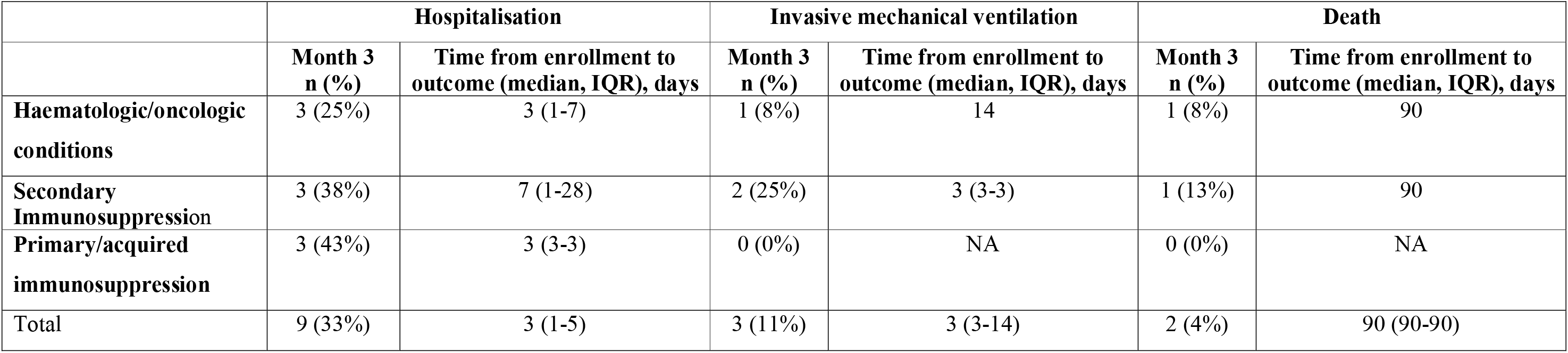
Clinical outcomes of participants at three months after enrolment.

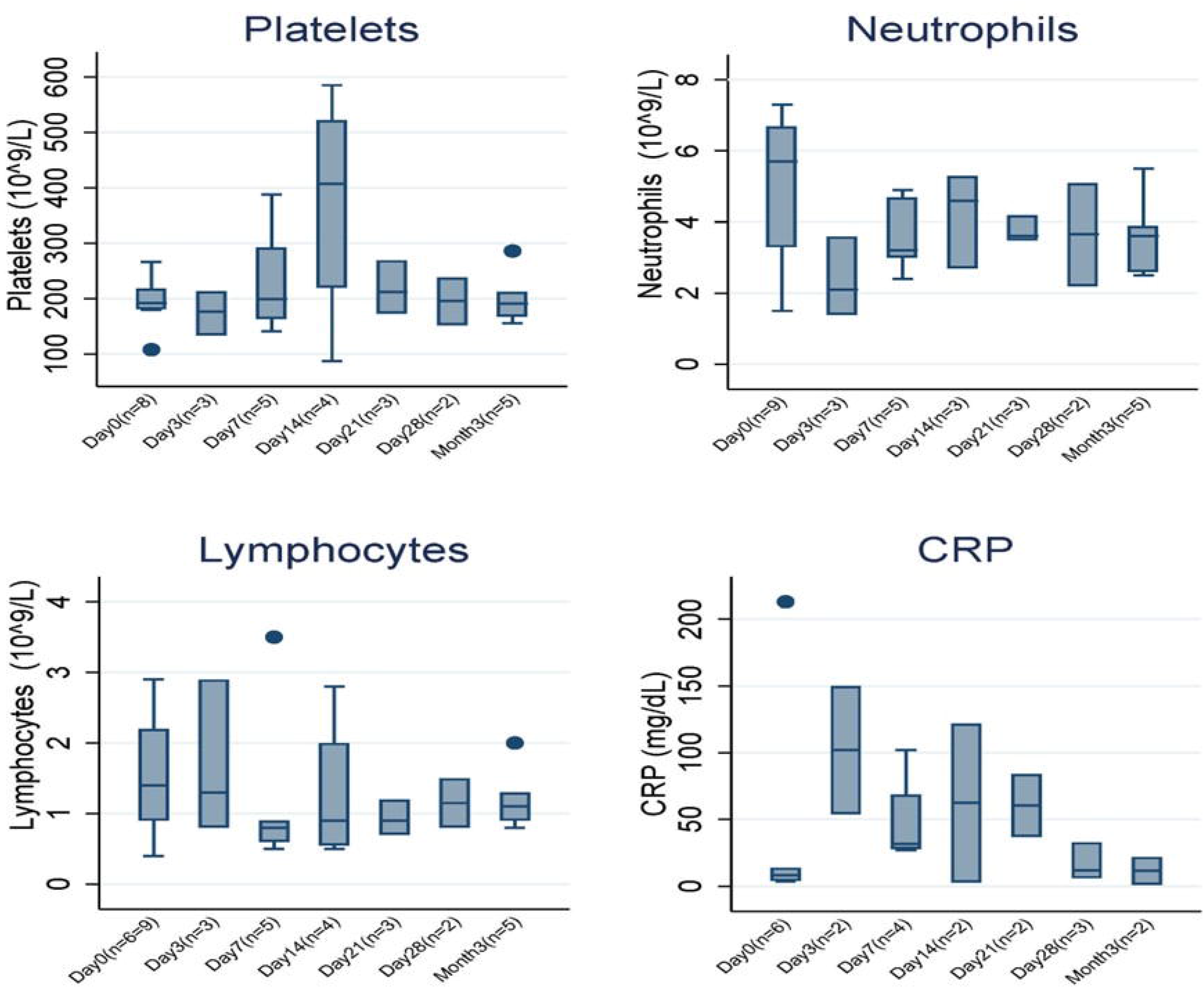

